# Factors associated with a willingness to accept latent tuberculosis infection treatment among non-U.S.–born individuals: a situational choice experiment

**DOI:** 10.64898/2026.09.09.26362665

**Authors:** Sutina Chou, Hélène E. Aschmann, Amy S. Tang, Meagan Lee, Zinnia Dong, Kit Lui, Yuqian Ouyang, Gina Chen, Katya L. Salcedo, Matthew Murrill, Mehabuba Rahman, Jennifer Flood, Andrew D. Kerkhoff, Priya B. Shete, Tracy Kuo Lin, the Tuberculosis Epidemiologic Studies Consortium

**Author notes:** contributed equally (Sutina Chou, Hélène E. Aschmann).

## Abstract

**Introduction:** Despite efforts to eliminate tuberculosis (TB) in the United States (U.S.), many individuals diagnosed with latent TB infection (LTBI) who are at increased risk of progressing to TB disease do not initiate TB preventive treatment.

**Methods:** Among adults receiving primary care in a federally qualified community health center in the San Francisco Bay Area, California, we conducted a cross-sectional online survey using a situational choice experiment to assess the circumstances in which individuals at risk for TB based on nativity would accept LTBI treatment. In 10 randomized choice tasks, participants answered whether or not they would accept LTBI treatment. In situations where LTBI treatment was accepted, participants were asked a follow-up question about whether they would change their response knowing that reinfection is still possible with LTBI treatment. Attributes assessed included risk level of progression to TB, side effects, changes to co-medication, cost, number of clinic visits, blood draws, and reinfection risk. We conducted a mixed-effects logistic regression to estimate factors associated with treatment acceptance and predicted acceptance across scenarios.

**Results:** Of 852 individuals who consented, 458 (53.8%) had complete responses and were included in the analysis. LTBI treatment was accepted in 69.4% of choice scenarios (3180/4580); among scenarios where treatment was initially accepted, 91.5% of scenarios (2911/3180) remained accepted when reinfection possibility was introduced. Compared to situations with low risk of TB progression, high TB progression risk increased the acceptance of LTBI treatment (adjusted odds ratio aOR 2.55, 95% Confidence Interval (CI) = 2.03–3.19). Compared with fatigue as a referent, risk of any liver injury/inflammation (aOR 0.29, 95% CI = 0.22–0.38), gastrointestinal side effects (aOR 0.53, 95% CI 0.41, 0.69), rash (aOR 0.62, 95% CI 0.48-0.80) and out-of-pocket cost of $100 (aOR 0.32, 95% CI = 0.26–0.41 compared to $0) were associated with lower acceptance of LTBI treatment.

**Conclusions:** The decision to accept LTBI treatment was sensitive to treatment characteristics, suggesting that shared decision-making should be broadly promoted among individuals diagnosed with or at risk of LTBI, particularly in diverse primary care settings serving individuals born in high TB burden countries, with attention to individual concerns such as cost and side effects to improve LTBI treatment uptake.

## Introduction/Background

Over 80% of tuberculosis (TB) cases in the United States (U.S.) likely represent progression of latent TB infection (LTBI).^1^ TB and LTBI disproportionally affect non-U.S.–born individuals, with LTBI prevalence as high as 31% in some settings.^2,3^ Given this reality, efforts to eliminate TB necessitate screening and providing treatment for LTBI. Despite LTBI being asymptomatic and non-infectious, individuals with LTBI harbor a 5% to 15% lifetime risk of progressing to TB disease if untreated.^4^

LTBI treatment decreases the risk of progression of LTBI to TB disease by 60-90%, and is a key pillar of the U.S. TB elimination strategy.^5,6^ Though effective treatment options exist (e.g. the U.S. Centers for Disease Control and Prevention (CDC) recommended short-course rifamycin-based regimens),^7–9^ initiation and completion rates of LTBI treatment remain low.^8,10–13^ While concerns about side effects, repeated clinic visits, blood draws, out-of-pocket costs, and drug-drug interactions represent significant barriers to LTBI treatment, the importance of these factors varies per individual.^14,12,15^ However, little is known about which factors patients value and whether their treatment preferences align with public health goals.

Previous research on patient preferences for LTBI treatment for individuals with LTBI is scarce and largely focused on high TB incidence settings.^14,16,17^ For example, a discrete choice experiment (DCE) study found that among people living in Uganda, treatment duration, pill burden, dose frequency, and dose adjustments of anti-retroviral therapy factored strongly into decision-making. In Canada, a low-TB-incidence setting, patients generally preferred regimens with higher effectiveness, fewer side effects, and shorter duration, but individual preferences were still substantially heterogeneous.^18^ However, this study did not focus on factors such as immigration status, international travel, or cost—an attribute particularly salient in non-universal coverage healthcare systems such as those in the United States.

Broadly, there remains limited evidence on how individuals at elevated risk of LTBI in the United States value trade-offs between treatment characteristics. To address this gap, we built on findings from a pilot study among non-U.S.–born individuals receiving primary care in San Francisco, which identified costs, drug-drug interactions, blood tests, and reinfection risk as factors influencing LTBI treatment decision-making.^12^

In this study, we evaluate the relative importance of these factors in influencing LTBI treatment acceptance. Leveraging a situational choice experiment (SCE) survey, an instrument based on decision science,^11^ we quantify how participants engage in trade-offs across treatment characteristics. By understanding the preferences of individuals at elevated risk of LTBI, we aim to generate evidence to inform clinical decision-making tools and strategies to improve acceptance, uptake, and completion of LTBI treatment.

## Methods

### Setting, participants, and recruitment process

In this cross-sectional observational study, we administered an online survey using a SCE among adult primary care patients at North East Medical Services (NEMS), a federally qualified community health center with over 13 clinic locations across the San Francisco Bay Area serving a predominantly Asian, non-U.S.–born population. Invited individuals were randomly sampled from patients who met all of the following eligibility criteria: (1) >=1 primary care visit at NEMS between October 2023 and October 2024 (2) age >=18 years, (3) birth in a country classified as having intermediate or high TB incidence based on World Health Organization criteria,^19^ (4) preferred language of English or Chinese (Mandarin and or Cantonese), (5) consented to be contacted by email or postal mail by their healthcare system, and (6) had an address on file. Patients who did not meet all the inclusion criteria were excluded. Prior LTBI testing or diagnosis was not required. We conducted recruitment through electronic mail (one batch of 43, followed by 18 batches of 499) and postal mail (one batch of 998), inviting individuals to complete an online survey hosted through the survey software Sawtooth (Lighthouse Studio version 9.16.2 by Sawtooth Software Inc., Provo, Utah, USA). Before starting the survey, individuals completed an online eligibility screener confirming that they were a NEMS patient 18 years or older born outside the United States and provided electronic consent. After completing the survey, participants received a $20 Target gift card as compensation for their time.

### Ethics

The study was approved by the University of California, San Francisco Institutional Review Board (reference #389568). Adhering to protocol,^18^ all eligible individuals received an informed consent question on the first page of the survey and individuals could decline or provide electronic consent to participate. Participants who provided informed consent were routed to the main survey. The CDC received aggregate data of the study for intervention evaluation. Program evaluation of aggregate survey data of the survey was reviewed by CDC, deemed not research, and conducted consistent with applicable federal law and CDC policy (see 45 C.F.R. part 46. 102 (l)).

### Survey administration

The surveys were designed and programmed in English, Simplified Chinese, and Traditional Chinese. Translation and quality checks were completed by NEMS staff (KL, YO, GC) with language proficiency to ensure accuracy across all versions. The surveys were self-administered; the recruitment letter that participants received contained a general QR code so they could asynchronously complete the survey on their own device (e.g. computer, smartphone, etc.). At the start of the SCE survey component, participants viewed a brief video featuring visual aids and audio narration explaining the SCE format. They were reminded that the situations presented were hypothetical and that they can either choose to accept or decline LTBI treatment in each scenario. After the survey instructions, there were a series of questions meant to facilitate information recall. If any comprehension questions were answered incorrectly, participants were prompted to select the correct response to move on.

### SCE Survey Instrument

SCEs are related to discrete choice experiments but present a single hypothetical scenario defined by multiple attributes.^12^ Participants are asked whether they would accept or decline the option described in each scenario. In the context of our study and many others in the healthcare field, SCE is a robust methodology for identifying and quantifying patient preferences in healthcare decision-making. This approach allows estimation of the relative importance of LTBI treatment attributes in decision-making.^20^

Attributes and levels were identified through a literature review of patient preferences and barriers to LTBI treatment. We then conducted a pilot study to confirm the most relevant barriers for this community and solicited stakeholder input by reviewing the results with clinical experts serving the eligible population.^21^ We also conducted language-concordant cognitive interviews with 8 primary care patients at NEMS to solicit feedback on their understanding of the survey tool, as well as on timing, wording, and instruction clarity. In response, we added additional instructions and a set of comprehension questions; additionally, we streamlined the follow-up question wording. However, we did not make any changes to the experimental design after conducting the pilot study or cognitive interviews. Additionally, we consolidated the number of levels per attribute to ensure a feasible sample size. We did not limit our design to exclude low-likelihood scenarios or potentially unrealistic hypothetical choice sets (e.g., monthly blood draws without any clinic visits), as our aim was to understand any potential cognitive dissonance among participants. No cognitive dissonance was identified during the cognitive interviews after pre-testing the initial survey.

Participants completed a randomly assigned set of 10 choice tasks from a software default of 300 possible scenarios,^22^ each presenting a single hypothetical scenario defined by six different attributes: 2-year risk of developing TB disease without LTBI treatment as low, medium, or high, one of a chosen list of side effects, changes to existing medication needed, out-of-pocket payment cost needed, clinic visits needed, and blood draws needed (for details, see **Table 1**). Default settings were used for all elements of survey design. Participants were asked, “Given the scenario above, would you accept preventive treatment?” with “yes” or “no” response options. For scenarios where participants initially accepted treatment, a follow-up question introduced potential reinfection risk by asking, “If you knew you could be reinfected with the TB germ when traveling to a high TB risk region in the future, what would you do?” with options to still accept LTBI treatment or reverse their decision and decline LTBI treatment. This conditional task was designed to assess the stability of treatment acceptance under additional risk information. An example choice task is shown in **Figure 1**, and the full example survey is provided in **Supplementary File 1.**

**Figure 1:**
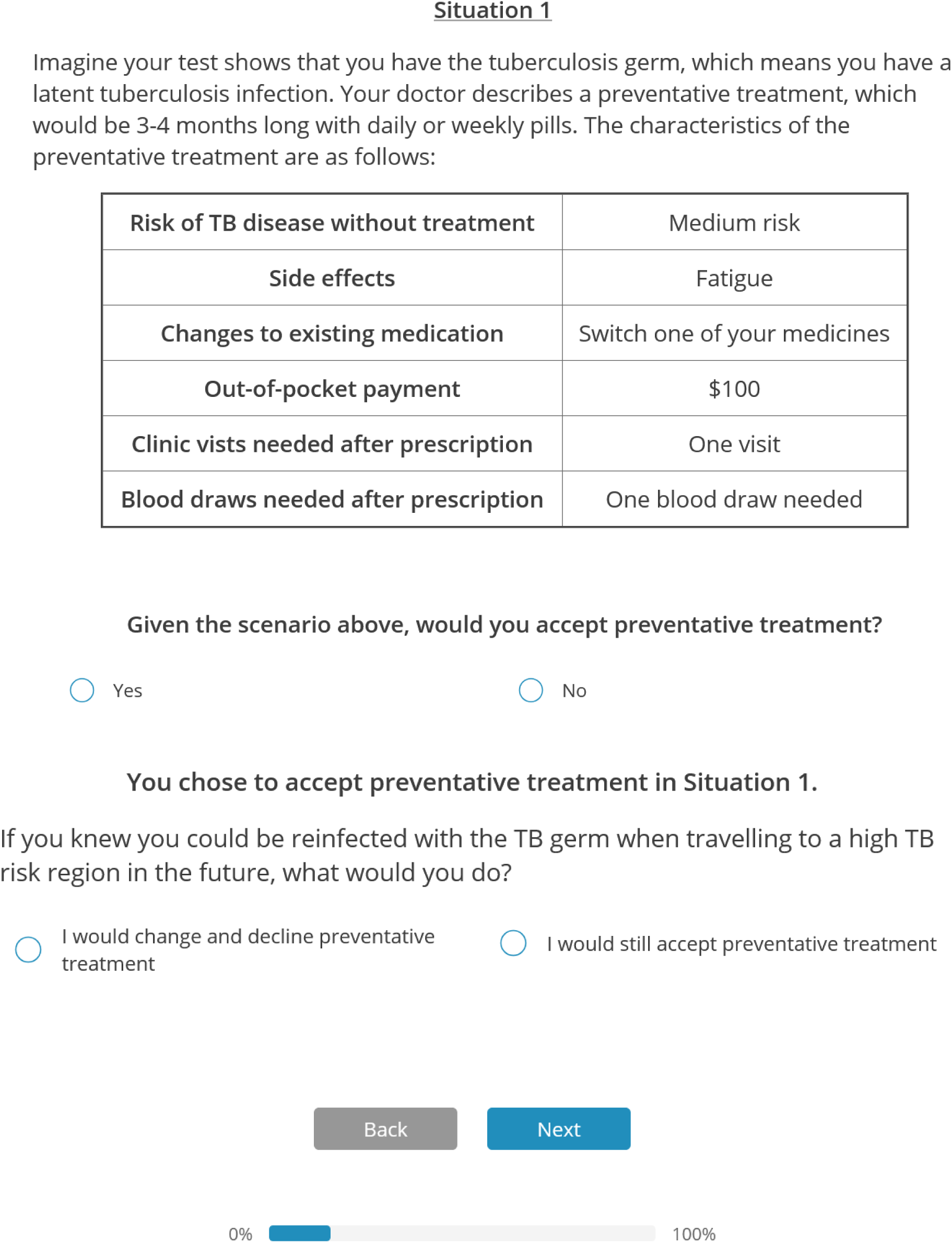
Example of a situation (and associated follow-up question) presented to participants

**Table 1:** Summary of attributes and levels used in survey.

| **Characteristic of preventive treatment** | **Possible levels for scenarios** |
| --- | --- |
| **Risk of TB disease without preventive treatment** | Low risk<br>Medium risk<br>High risk |
| **Potential side effects** | Skin rash<br>Fatigue<br>Gastrointestinal inflammation<br>Liver injury or inflammation |
| **Changes to existing medication needed** | No change needed<br>Stop a medicine for 4 months<br>Switch one of your medicines for 4 months |
| **Out-of-pocket payment cost needed** | \$0<br>\$50<br>\$100 |
| **Clinic visits needed** | 0 visits<br>1 visit<br>4 visits |
| **Blood draws needed** | No blood draws needed<br>One blood draw needed<br>Monthly blood draws needed |
| **If LTBI treatment accepted:** |  |
| **Reinfection risk** | Yes<br>No |

### Sample size calculation

We estimated the sample size using the formula n > 10k/p, where n is the sample size, k is the number of non-zero parameters to be estimated, and p is the probability of the least frequently chosen alternative.^23^ With 14 parameters in the final design, and assuming no LTBI treatment would be chosen in 10% of scenarios, this required at least 1400 observations per subgroup (three age groups of 18-39, 40-59, and 60+), corresponding to 140 participants per subgroup with 10 choice tasks per participants, totaling 420 across the three age groups. Simulation analyses indicated that a sample of 400 participants would yield predicted acceptance probabilities of choosing LTBI treatment ranging from 16 to 89% across different scenarios with a precision of +/-2.5 to 4.5%. Thus, we aimed to enroll 400-500 participants, with analysis limited to main effects only.

### Data cleaning

We performed data cleaning to include only complete responses. We excluded those who did not complete the survey and participants with a completion time of less than 3 minutes. We determined this threshold through pilot testing to ensure actual engagement, as the survey actually required approximately 15 minutes to complete.

### SCE Analysis

We conducted a mixed-effects logistic regression (using the function glmer in the lme4 R package^24^) that included all attribute levels as fixed effects and a random intercept for individual participants, as we expected choices to be correlated within participants across scenarios. We calculated adjusted odds ratios (aORs) for all levels relative to the reference level. In addition, we estimated difference in the predicted probability of accepting LTBI treatment for each attribute level compared with the reference level. Predicted probabilities of accepting LTBI treatment were obtained across all possible scenarios for all participants. We then calculated the mean predicted probabilities for all scenarios that contained a specific attribute level and for the corresponding reference level. Finally, we calculated the differences between these marginal probabilities, which represent absolute changes in predicted proportion of participants accepting LTBI treatment. We reported 95% confidence interval using the 2.5^th^ and 97.5^th^ percentiles of the differences. We also conducted subgroup analysis for three age groups: ages 18-39, 40-59, and 60+. This resulted in four different models, one for each age group and one for all ages. The results for each model are reported in **Supplemental Table 3**.

### Willingness to pay analysis

We calculated the willingness to pay (WTP) for each of the treatment characteristics (i.e., attribute levels) using the following formula: WTP = (preference weight of attribute ÷ preference weight of out-of-pocket cost) x out-of-pocket cost, where weight refers to the coefficient for each variable in the logistic regression. WTP represents the monetary value participants place on receiving or avoiding the given attribute level relative to the reference level. The results can be interpreted as the amount someone is willing to pay to receive/avoid a characteristic relative to each category’s reference group. We calculated WTP across all non-reference attribute levels for out-of-pocket costs of $50 and $100 (see **Table 2**). Positive values indicate the amount participants would pay to avoid an attribute level relative to the reference level (i.e., the level is less preferred than the reference). Negative values indicate the amount participants would need to be compensated to give up an attribute level in favor of the reference level (i.e., the level is more preferred than the reference).

**Table 2:** Willingness to pay (WTP) calculated for all attribute levels compared to the preference weight for $100 and $50 out-of-pocket costs, respectively. (SE: standard error, CI: confidence interval, USD: US Dollars)

| | Coefficient<br>(SE) | Compared to \$100 out-of-pocket cost | | Compared to \$50 out-of-pocket cost | |
| --- | --- | --- | --- | --- | --- |
|  |  | WTP<br>(USD) | [95% CI] | WTP<br>(USD) | [95% CI] |
| TB risk (reference: low) |  |  |  |  |  |
| medium | 0.67 (0.11) | -59.25 | [-82.16, -36.33] | -47.14 | [-68.81, -25.48] |
| high | 0.93 (0.12) | -82.86 | [-109.09, -56.63] | -65.93 | [-92.45, -39.41] |
| Side effects (reference: fatigue) |  |  |  |  |  |
| Skin rash | -0.48 (0.13) | 42.60 | [17.80, 67.40] | 33.90 | [12.44, 55.35] |
| Gastrointestinal inflammation | -0.64 (0.13) | 56.56 | [30.59, 82.52] | 45.00 | [21.51, 68.49] |
| Liver injury or inflammation | -1.24 (0.13) | 110.29 | [77.82, 142.77] | 87.75 | [53.96, 121.55] |
| Drug-drug interactions<br>(reference: none) |  |  |  |  |  |
| Switch other medication | -0.33 (0.11) | 29.39 | [8.67, 50.11] | 23.38 | [5.90, 40.86] |
| Stop other medication | -0.44 (0.11) | 39.18 | [17.81, 60.56] | 31.17 | [12.49, 49.86] |
| Clinic visits (reference: 0) |  |  |  |  |  |
| 1 | 0.18 (0.11) | -16.08 | [-36.10, 3.94] | -12.79 | [-29.04, 3.45] |
| 4 | -0.19 (0.11) | 17.08 | [-2.75, 36.91] | 13.59 | [-2.54, 29.72] |
| Blood draws (reference: 0) |  |  |  |  |  |
| 1 | -0.14 (0.11) | 12.77 | [-7.13, 32.67] | 10.16 | [-5.87, 26.20] |
| monthly | -0.29 (0.11) | 25.58 | [5.26, 45.89] | 20.35 | [3.41, 37.29] |

**Note:** Willingness to pay (WTP) was calculated using the following formula: (preference weight of attribute level ÷ preference weight of out-of-pocket cost) x out-of-pocket cost, where preference weight refers to the coefficient for each variable in the logistic regression (see first column). The coefficients were -0.71 (0.12) for \$50 and -1.13 (0.12) for \$100. Attributes with positive coefficients indicate that LTBI treatment was more preferred; for example, scenarios with high TB risk increase LTBI treatment acceptance. Attributes with negative coefficients indicate that LTBI treatment was less preferred with the feature compared to the reference level (e.g. liver injury is less preferred than the baseline of fatigue).
Positive WTP values indicate the amount that participants were willing to pay to avoid the attribute level. For example, participants would be willing to pay \$42.60 to avoid skin rash as a side effect relative to the baseline of fatigue. Negative WTP values represent the amount that would be required to compensate participants for giving up attribute levels that increase LTBI treatment acceptance. For example, participants were more willing to accept LTBI treatment at higher risk of TB compared to low risk; LTBI treatment was worth \$82.86 under conditions of high TB risk and \$59.25 under conditions of medium TB risk, compared to low risk.

### Sensitivity Analysis

We conducted two sensitivity analyses. First, we conducted a mixed-effects logistic regression that included reinfection risk as a variable. For each choice task, we incorporated the follow-up response reflecting risk of reinfection. For scenarios in which participants initially declined treatment and were not presented with the reinfection follow-up, we assumed the decision would remain unchanged and LTBI treatment would still be declined, as the situation favored LTBI treatment less and would not increase the likelihood of accepting treatment. Second, we conducted a sensitivity analysis that included participants who had only partially completed the survey. All analyses were conducted in R version 4.1.2.

## Results

We sent 9,025 emails and 998 postal mail letters to invite eligible primary care patients at NEMS to participate in the survey, for a total of 10,023 individuals invited. Patients that were invited received either an email or a mail letter, but not both. Of the 10,023 invited individuals, 852 people consented (a response rate of 8.5%); from this group, after the data cleaning process described above, 458 participants were included in the main analysis. For more information on the flow of individuals from invitation to main analysis, please see **Supplemental Figure 1**. **Supplemental Table 1** compares individuals who were invited and responses from participants who were ultimately included in the main analysis. It also provides information about the age distribution and language preferences of both groups. Compared with the invited population, more individuals of age 18-39 years and fewer individuals with English as their primary language participated in the survey.

### SCE Analysis

**Figure 2** presents the adjusted odds ratios (aOR) corresponding to each attribute level, along with the difference in the predicted proportion of individuals accepting LTBI treatment. Compared to reference scenarios in which risk of progression to TB disease was low, being at medium risk and high risk of progression to TB disease without LTBI treatment were associated with a 1.95 (95% CI 1.57–2.43) times and 2.55 (95% CI 2.03–3.19) times higher odds of preferring treatment, respectively. Among other factors related to LTBI treatment, risk of liver injury or inflammation (compared to fatigue) as a treatment side effect was associated with the least preference for LTBI treatment (aOR 0.29, 95% CI 0.22–0.38), followed by $100 out-of-pocket cost compared to $0 (aOR 0.32, 95% CI 0.26–0.41).

**Figure 2:**
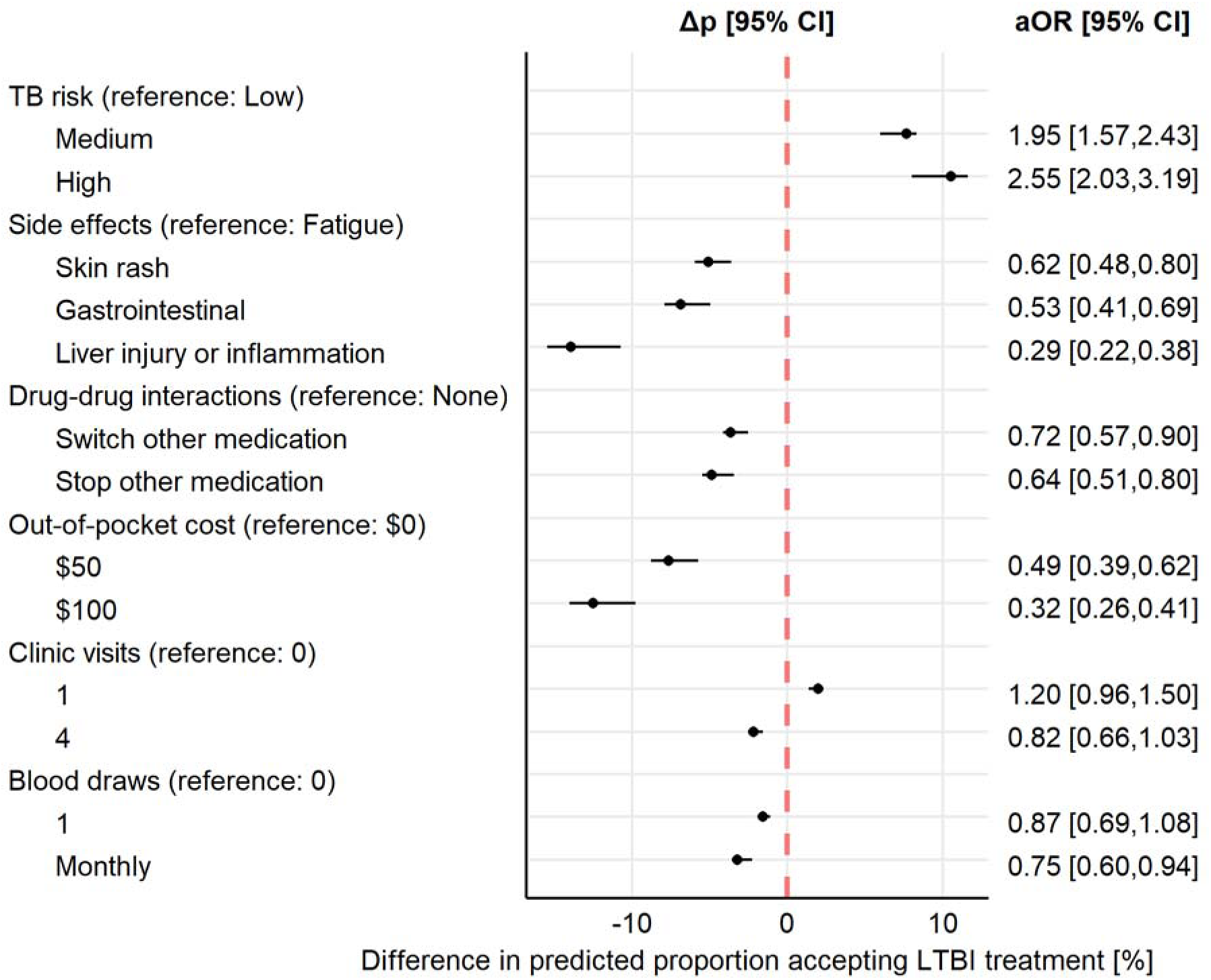
Association of factors with preference for latent tuberculosis (TB) infection (LTBI) treatment over no treatment (N=458). Adjusted odds ratios (aOR) were calculated using mixed effects logistic regression with fixed effects for all factors in the model simultaneously and random effects for individual participants. The difference in the predicted proportion (Δp) of individuals who would prefer LTBI treatment over no treatment was calculated for all combination of scenarios, and we compared the effects of different factor levels (e.g., high vs low risk of TB) across scenarios. TB risk was explained as a two-year risk without LTBI treatment, where the risk is much lower with LTBI treatment. Potential side effects were described as fatigue, skin rash, gastrointestinal inflammation, and liver injury or inflammation was described as reversible by stopping LTBI treatment. Clinic visits and blood draws were described as after treatment initiation to check liver and kidney function. aOR: adjusted odds ratio, CI: confidence interval, LTBI: latent tuberculosis infection, TB: tuberculosis, Δp: difference in predicted proportion accepting LTBI treatment

The predicted proportion of individuals accepting LTBI treatment ranged from 43.0% to 89.8%, depending on the scenario (see **Supplemental Table 2**). LTBI treatment was accepted in 69.4% of scenarios; among scenarios where treatment was initially accepted, 91.5% of scenarios remained accepted when reinfection risk was introduced. Across scenarios, a medium risk of TB increased the predicted proportion of individuals accepting LTBI treatment by +7.7% (95% CI 6.0–8.3%) and a high risk by +10.5% (95% CI 8.0–11.6%) (**Figure 2**). Conversely, the risk of liver injury or inflammation decreased the proportion accepting LTBI treatment by -13.9% (95% CI -15.4 – -10.7%) and the $100 out-of-pocket cost by -12.5% (95% CI -14.0 – -9.80%).

In a sensitivity analysis that included reinfection risk, the results for all other attribute levels remained similar to the main analysis. However, risk of reinfection was associated with a 0.58 (95% CI 0.51–0.65) times reduced odds of preferring to take LTBI treatment (**Supplemental Figure 2**). In a second sensitivity analysis, we included 32 participants who did not complete all choice tasks; results did not differ meaningfully from the main analysis (**Supplemental Figure 3**).

In a subgroup analysis for each age group (i.e. 18-39, 40-59, 60+), high TB risk level universally increased the odds of accepting LTBI treatment (**Supplemental Table 3**). Similarly, liver injury or inflammation and out-of-pocket costs reduced the odds of accepting LTBI treatment across all age groups. Stopping another medication and monthly blood draws were significant only in the group of patients aged 60+ years, among whom they reduced the odds of accepting LTBI treatment by aOR 0.43 (95% CI 0.29–0.65) and aOR 0.57 (95% CI 0.38–0.84).

### Willingness-to-pay analysis

**Table 2** presents willingness-to-pay (WTP) estimates across all attributes and levels at $50 and $100 out-of-pocket cost levels. These results quantify the average value in USD that participants ascribed to an attribute level compared to the reference level. Per expectations, increased TB risk is associated with increased LTBI treatment acceptability. At $100 out-of-pocket cost and compared to low risk of TB, LTBI treatment under conditions of medium TB risk was worth $59 (95% CI $36-$82) to participants, whereas treatment under conditions of high risk was worth $83 (95% CI $56-$109). At both out-of-pocket cost levels, participants demonstrated the same pattern in willingness to pay to avoid side effects relative to fatigue; for example, participants were willing to pay $110 (95%CI $78-$143) to avoid risk of liver injury (at $100 out-of-pocket cost). Overall, participants were willing to pay the most to avoid liver injury or inflammation, followed by gastrointestinal discomfort, skin rash, and fatigue. Participants were also willing to pay to avoid changes to concomitant medications as part of LTBI treatment. At $100 out-of-pocket cost, participants were willing to pay $29 (95% CI $9-$50) to avoid switching a medication and $39 (95% CI $18-$61) to avoid stopping a medication. WTP analysis results for the clinic visits attribute at one visit or four visits compared to no clinic visits were not statistically significant. For the blood draw attribute, compared to no blood draws, participants were willing to pay $26 (95% CI $5-$46) to avoid monthly blood draws at $100 out-of-pocket cost; however, there was no significant avoidance for a single blood draw.

## Discussion

Our study shows that LTBI treatment acceptance is sensitive to an individual’s preferences. Most attributes were relevant to participants’ decision-making; however, risk of TB disease, side effects, out-of-pocket costs, and reinfection possibility had the largest effects on LTBI treatment acceptance. The observed direction of the effects from these attributes was as expected; higher risk of TB disease was associated with increased LTBI treatment acceptance. Additionally, more severe side effects, higher out-of-pocket costs, greater treatment burden (e.g. drug-drug interactions, clinic visits, and blood draws), and reinfection were associated with lower LTBI treatment acceptance. Scenarios that assumed a medium or high risk of TB disease were associated with approximately twofold higher odds of accepting LTBI treatment, underscoring the importance of risk communication when considering approaches to shared decision making for LTBI treatment.

Predicted LTBI treatment acceptance ranged from 43 to 90% across scenarios, consistent with historical rates from NEMS, where a 2019 cascade found a treatment prescription rate of approximately 59% among adult primary care patients who were seen for regular or preventative medical visits and tested positive for TB.^25^ However, treatment initiation rates as low as 10% have been reported amongst individuals with LTBI in the United States.^26^ A preference study in Canada among a similar population found that 75% accepted LTBI treatment, although the highest predicted proportion was 86%, which may be higher because treatment would be no-cost in Canada.^18,27^ Preference studies from high-incidence settings have shown that risk awareness increased LTBI treatment acceptance, and importantly, that TB education helps reduce barriers to LTBI treatment uptake.^28,29^ Our estimates may therefore be conservative, as participants only received broad educational content about TB as opposed to specific TB risk education. Overall, LTBI treatment among non-U.S.–born individuals in the United States appears to be a preference-sensitive decision that can be modified with educational interventions and clinical shared-decision making in TB prevention.^30^

Our results have implications for helping improve person-centered service delivery for LTBI treatment. Out-of-pocket costs had a particularly large impact on LTBI treatment acceptance, consistent with our pilot survey findings.^21^ Even when treatment is provided at no cost, indirect costs (e.g., travel, lost wages) may reduce acceptance, suggesting that financial support or incentives could improve acceptance and uptake—consistent with prior literature.^31,32^ Another important factor was the side effect of liver injury, which could be addressed by offering rifamycin-based (as opposed to isoniazid-based) LTBI treatments along with careful medication counselling. However, all regimens have benefits and drawbacks; although rifamycin-based LTBI treatment carries less potential for liver injury, there are other risks such as drug-drug interactions. This concern is particularly relevant for older populations, who are less likely to accept LTBI treatment if it requires medication changes while also being more likely to be on other medications that could interact with LTBI treatment.^30^ Together, these findings highlight the need for tailored LTBI treatment strategy that align regimen safety, toxicity profiles, and patient preferences while minimizing drug-drug interactions.

### Limitations

The probability of progressing to TB disease and experiencing side effects was not specified as a sliding scale number in this study because our pilot study showed that it was too cognitively burdensome for participants who had limited statistical literacy. Quantitative risk information was not well understood in pilot testing, even with visual aids; as such, we decided against presenting probabilities.^12,13^ As a result, we could not assess how participants perceived the risk of TB progression. We also did not specify what low, medium, or high risk of progression to TB meant in terms of numbers. Thus, the measured effect reflects how participants interpret qualitative risk description.

On the same note, SCE are an inherently hypothetical methodology, meaning that participants may have answered questions about situations that they are not or may not ever be in (i.e. someone exhibits preference against stopping medications but does not actually take any medications). Similarly, it is possible that social desirability, acceptability bias and other factors related to patient comfort level with NEMS clinical visits may have affected our results.

Both limitations have to do with how representative the SCE scenario is to a participant’s current circumstances. However, SCE as a methodology is meant to be hypothetical in order to shed light specifically on a participant’s internal decision-making logic and is not intended to reflect their literal situation. The methodology offers two key insights: it can quantify (1) the importance of certain attributes (i.e. side effects vs. blood draws) and (2) within attributes, the importance of certain levels (i.e. one clinic visit vs. four). However, future research can address these limitations by collecting additional participant information (i.e. medication regimen) and providing further specification (i.e. clarifying where care would take place and with whom).

Additionally, participants received recruitment emails and survey materials in their preferred written language (English, Traditional Chinese, or Simplified Chinese) and were directed to a corresponding survey with an instructional video. While written materials always matched participants’ stated language preference, the audio voiceover may not have aligned with all participants’ preferred spoken language (e.g., Mandarin vs. Cantonese), which may have affected comprehension; however, all content was also presented in text.

Furthermore, our data comes from a single community health center with a largely homogenous population – NEMS serves a predominantly Asian, non-U.S.–born population. As such, our sample is not representative of the general non-U.S.–born population. Along similar lines, it is possible that our recruitment strategies affected the distribution of participants across age groups. For additional information on age breakdown of invited individuals versus complete survey responses, see **Supplemental Table 1**. For example, older adults may be less likely to use email, and even if they received a letter, may be less likely to type the survey link into a browser. This could have led to an increase in younger participants and decrease in older participants in the analyzed group compared to those that were invited as well as compared to the broader non-U.S.–born population. The age-based selection bias in those who did not fill out our survey versus those who did may also have affected our results.

## Conclusion

Among our sample of non-U.S.–born individuals, LTBI treatment acceptance varied depending on their perceived risk level of TB disease progression, overall cost of care, side effects, reinfection possibility, potential medication changes, and blood draws required. By quantifying and evaluating individual preferences, we have generated evidence supporting the need for shared decision-making around LTBI treatment initiation and the attributes that shared decision-making tools should address. Our data can be used to inform the design and implementation of strategies to improve the provision and uptake of LTBI treatment such as financial accessibility. In particular, regimen selection should be attentive to overall patient characteristics by considering both potential side effects, including liver injury risk and possible drug-drug interactions. Even under less favorable conditions, many individuals were willing to accept treatment, supporting broader engagement in LTBI care and shared decision making.

## Data Availability

All data produced in the present study are available upon reasonable request to the authors.

## Funding

This work was supported by the Tuberculosis Epidemiologic Studies Consortium III, sponsored by the Centers for Disease Control and Prevention (Award 75D30121C12879). HEA was supported by a Postdoc Mobility fellowship (214129) from the Swiss National Science Foundation, and by the UCSF Center for Tuberculosis, NIH/NIAID P30: TB Research Advancement Center (UC TRAC) P30AI168440, and NIH/NIAID R25: TB Research and Mentorship Program (TB RAMP) 1R25AI147375

## Acknowledgments

We thank the UCSF-Bay Area Center for AIDS Research (CFAR) for providing a Sawtooth license (P30 AI027763). This publication was supported by the National Center for Advancing Translational Sciences, National Institutes of Health, through UCSF-CTSI

Grant Number UL1 TR001872. The findings and conclusions are those of the authors and do not necessarily represent the views of the federal government. References in this article to any specific commercial products, process, service, manufacturer, or company do not constitute its endorsement or recommendation by the U.S. government, the CDC, the California Department of Public Health, or the California Health and Human Services Agency. Most importantly, we thank participants in our study who shared their time and perspectives to inform TB prevention.

## COI

The authors declare no conflicts of interest.

## Abbreviations

TB: Tuberculosis
U.S.: United States
LTBI: Latent Tuberculosis Infection
LTBI Treatment: Latent Tuberculosis Infection Preventive Treatment
SCE: Situational Choice Experiment
CDC: Centers for Disease Control and Prevention
DCE: Discrete Choice Experiment
NEMS: North East Medical Services
aORs: Adjusted odds ratios
WTP: Willingness-to-pay
CI: Confidence interval

## Disclaimer

The findings and conclusions in this article are those of the authors and do not necessarily represent the views or opinions of the California Department of Public Health, the California Health and Human Services Agency, the Centers for Disease Control and Prevention, or the National Institutes of Health.

## Appendix: Supplemental Materials

### Files

**Supplementary File 1:** Full example survey script

### Figures

**Supplemental Figure 1:** Study flow chart. Invitations to the online survey were sent by email and postal mail.

**Supplemental Figure 1:** Sensitivity analysis including reinfection risk. This analysis shows the association of all factors with preference for LTBI treatment over no LTBI treatment. Adjusted odds ratios (aOR) were calculated using mixed effects logistic regression with fixed effects for all factors and random effects for individual participants. The difference in the predicted proportion of individuals who would prefer LTBI treatment over no LTBI treatment was calculated for all combination of scenarios, and we compared the effects of different factor levels (e.g., high vs low risk of tuberculosis) across scenarios. An aOR above 1 means LTBI treatment is more likely to be accepted with this feature (attribute level) compared to the reference.

aOR: adjusted odds ratio, CI: confidence interval, TB: tuberculosis, LTBI: latent tuberculosis infection, Δp: difference in predicted proportion accepting LTBI treatment

**Supplemental Figure 2:** Sensitivity analysis including partial responses (N=490). This analysis shows the association of all factors with preference for LTBI treatment over no LTBI treatment. Adjusted odds ratios (aORs) were calculated using mixed effects logistic regression with fixed effects for all factors and random effects for individual participants. The difference in the predicted proportion of individuals who would prefer LTBI treatment over no LTBI treatment was calculated for all combination of scenarios, and we compared the effects of different factor levels (e.g., high vs low risk of tuberculosis) across scenarios. An aOR above 1 means LTBI treatment is more likely to be accepted with this feature (attribute level) compared to the reference.

aOR: adjusted odds ratio, CI: confidence interval, TB: tuberculosis, LTBI: latent tuberculosis infection, Δp: difference in predicted proportion accepting LTBI treatment

### Tables

**Supplemental Table 1:** Comparison of age and survey language of invited individuals and individuals who were included in the main analysis. Responses analyzed came from participants who opened the survey, completed the eligibility screener and were eligible, consented, responded to all questions, and completed the survey in 3 or more minutes. We did not include sex as a variable in the survey and therefore only know the sex distribution for the invited population.

**Supplemental Table 2:** Scenarios minimizing and maximizing the predicted acceptance rate, i.e., the predicted proportion of individuals who prefer LTBI treatment over no LTBI treatment.

TB: tuberculosis, LTBI: latent tuberculosis infection

**Supplemental Table 3:** Subgroup analysis by age: association of factors with preference for LTBI treatment over no LTBI treatment. The analysis used mixed effects logistic regression (all fixed effects except for random intercept by participant). An OR above 1 means LTBI treatment is more likely to be accepted with this feature (attribute level) compared to the reference. The results are reported for four models: one for each age group, and one for all ages.

aOR: adjusted odds ratio, CI: confidence interval, TB: tuberculosis, LTBI: latent tuberculosis infection

## Notes

### Competing Interest Statement

The authors have declared no competing interest.

### Author Declarations

The Institutional Review Board of the University of California, San Francisco gave ethical approval for this work (reference #389568).

## Bibliography

1. Shea KM, Kammerer JS, Winston CA, et al. Estimated Rate of Reactivation of Latent Tuberculosis Infection in the United States, Overall and by Population Subgroup. American Journal of Epidemiology 2014; 179: 216–225.

2. Yuen CM, Kammerer JS, Marks K, et al. Recent Transmission of Tuberculosis — United States, 2011–2014. PLOS ONE 2016; 11: e0153728.

3. Campbell JR, Winters N, Menzies D. Absolute risk of tuberculosis among untreated populations with a positive tuberculin skin test or interferon-gamma release assay result: systematic review and meta-analysis. BMJ 2020; 368: m549.

4. Kiazyk S, Ball T. Latent tuberculosis infection: An overview. Can Commun Dis Rep 2017; 43: 62–66.

5. Price C, Nguyen AD. Latent Tuberculosis. In: StatPearls. Treasure Island (FL): StatPearls Publishing, http://www.ncbi.nlm.nih.gov/books/NBK599527/ (2026, accessed 1 July 2026).

6. CDC. Tuberculosis Elimination Priorities. National Center for HIV, Viral Hepatitis, STD, and Tuberculosis Prevention, https://www.cdc.gov/nchhstp/priorities/tuberculosis-elimination.html (2024, accessed 7 January 2026).

7. Zenner D, Beer N, Harris RJ, et al. Treatment of Latent Tuberculosis Infection: An Updated Network Meta-analysis. Ann Intern Med 2017; 167: 248–255.

8. Campbell JR, Trajman A, Cook VJ, et al. Adverse events in adults with latent tuberculosis infection receiving daily rifampicin or isoniazid: post-hoc safety analysis of two randomised controlled trials. The Lancet Infectious Diseases 2020; 20: 318–329.

9. Sterling TR, Njie G, Zenner D, et al. Guidelines for the treatment of latent tuberculosis infection: Recommendations from the National Tuberculosis Controllers Association and CDC, 2020. American Journal of Transplantation 2020; 20: 1196–1206.

10. Holzman SB, Perry A, Saleeb P, et al. Evaluation of the Latent Tuberculosis Care Cascade Among Public Health Clinics in the United States. Clin Infect Dis 2022; 75: 1792–1799.

11. Vonnahme LA, Raykin J, Jones M, et al. Using Electronic Health Record Data to Measure the Latent Tuberculosis Infection Care Cascade in Safety-Net Primary Care Clinics. AJPM Focus 2023; 2: 100148.

12. Aschmann HE, Tang A, Lee M, et al. Preferences for Tuberculosis Preventive Therapy in Primary Care Settings Among At-risk Immigrant Communities. In: 2024 Annual Meeting of the Society of General Internal Medicine. Springer, p. S798.

13. Shete PB, Murrill MT, Tatum KM, et al. Identifying Gaps in Tuberculosis Preventive Care for Non-U.S.-born Persons at Community Health Clinics in the United States. Open Forum Infectious Diseases 2025; 12: ofaf589.

14. Aschmann HE, Musinguzi A, Kadota JL, et al. Preferences of people living with HIV for features of tuberculosis preventive treatment regimens in Uganda: a discrete choice experiment. J Int AIDS Soc 2024; 27: e26390.

15. Vermeulen M, Scarsi KK, Furl R, et al. Patient and provider preferences for long-acting TB preventive therapy. IJTLD Open 2025; 2: 276–283.

16. Hirsch-Moverman Y, Strauss M, George G, et al. Paediatric tuberculosis preventive treatment preferences among HIV-positive children, caregivers and healthcare providers in Eswatini: a discrete choice experiment. BMJ Open 2021; 11: e048443.

17. Strauss M, Wademan DT, Mcinziba A, et al. TB preventive therapy preferences among children and adolescents. Int J Tuberc Lung Dis 2023; 27: 520–529.

18. Mohammadi T, Bansback N, Marra F, et al. Testing the External Validity of a Discrete Choice Experiment Method: An Application to Latent Tuberculosis Infection Treatment. Value in Health 2017; 20: 969–975.

19. World Health Organization. Global Tuberculosis Report 2025, https://www.who.int/teams/global-programme-on-tuberculosis-and-lung-health/tb-reports/global-tuberculosis-report-2025 (12 November 2025, accessed 9 January 2026).

20. Chrzan K. Situational Choice Experiments for Marketing Research: How to Design, Analyze and Report Them. Sawtooth Software, Inc., https://content.sawtoothsoftware.com/assets/c6112de1-d968-4754-a271-fab97555e831 (November 2022).

21. Abstracts from the 2024 Annual Meeting of the Society of General Internal Medicine. J GEN INTERN MED 2024; 39: 137–1008.

22. Sawtooth Software, Inc. The CBC System for Choice-Based Conjoint Analysis. Sawtooth Software, Inc., https://sawtoothsoftware.com/resources/technical-papers/cbc-technical-paper (August 2017, accessed 31 March 2025).

23. Peduzzi P, Concato J, Kemper E, et al. A simulation study of the number of events per variable in logistic regression analysis. J Clin Epidemiol 1996; 49: 1373–1379.

24. Bates D, Mächler M, Bolker B, et al. Fitting Linear Mixed-Effects Models Using lme4. J Stat Soft; 67. Epub ahead of print 2015. DOI: 10.18637/jss.v067.i01.

25. Tang AS, Mochizuki T, Dong Z, et al. Can Primary Care Drive Tuberculosis Elimination? Increasing Latent Tuberculosis Infection Testing and Treatment Initiation at a Community Health Center with a Large Non-U.S.-born Population. J Immigrant Minority Health 2023; 25: 803–815.

26. Mancuso JD, Miramontes R, Winston CA, et al. Self-reported Engagement in Care among U.S. Residents with Latent Tuberculosis Infection: 2011–2012. Ann Am Thorac Soc 2021; 18: 1669–1676.

27. Guo N, Marra CA, FitzGerald JM, et al. Patient Preference for Latent Tuberculosis Infection Preventive Treatment: A Discrete Choice Experiment. Value in Health 2011; 14: 937–943.

28. Aschmann HE, Musinguzi A, Kadota JL, et al. Preferences of people living with HIV for features of tuberculosis preventive treatment regimens – a discrete choice experiment. medRxiv 2023; 2023.09.13.23295043.

29. Musinguzi A, Aschmann HE, Kadota JL, et al. Preference for daily (1HP) vs. weekly (3HP) isoniazid-rifapentine among people living with HIV in Uganda. IJTLD OPEN 2024; 1: 83– 89.

30. Boyd CM, Darer J, Boult C, et al. Clinical Practice Guidelines and Quality of Care for Older Patients With Multiple Comorbid DiseasesImplications for Pay for Performance. JAMA 2005; 294: 716–724.

31. Rocha C, Montoya R, Zevallos K, et al. The Innovative Socio-economic Interventions Against Tuberculosis (ISIAT) project: an operational assessment. The International Journal of Tuberculosis and Lung Disease 2011; 15: S50–S57.

32. Barss L, Moayedi-Nia S, Campbell JR, et al. Interventions to reduce losses in the cascade of care for latent tuberculosis: a systematic review and meta-analysis. The International Journal of Tuberculosis and Lung Disease 2020; 24: 100–109.

