## Appendix for "Factors associated with a willingness to accept latent tuberculosis infection treatment among non-U.S.–born individuals: a situational choice experiment"

**Supplemental File 1:** Full example survey script

**Study Eligibility**

Are you a NEMS patient? (Yes/No)

As of today, how old are you? (numeric response)

Were you born in the United States? (Yes/No)

**If ineligible to participate (not NEMS patient, under 18, or U.S-born)**

Based on your responses, you are ineligible to participate in this survey. Thank you for your interest.

**Consent**

**Study information**

We are asking you to take part in a research study. NEMS is participating in this research study with the University of California, San Francisco (UCSF). The study is led by Dr. Tracy Lin at UCSF and Dr. Amy Tang at NEMS. In this study, the researchers are doing a survey to learn more about your perspective on preventive treatment for latent tuberculosis infection. The CDC (Centers for Disease Control and Prevention) is paying for this research. About 500 people will participate in different surveys in this study. Being in this study is optional, meaning that you do not have to take part.

What will happen if I take part in this study?

If you choose to be in the study, you will complete a survey. This survey will capture your perspective on preventative treatment for latent tuberculosis infection through a series of hypothetical questions. People who have latent tuberculosis are healthy and have no symptoms, but they are at risk of developing active tuberculosis, a serious illness. This survey will help us learn more about when people should be tested and treated for latent tuberculosis. The survey will take about 10-20 minutes to complete. Some of the survey questions may make you feel uncomfortable or raise unpleasant memories. You can skip most questions that you do not want to answer or stop the survey at any time.

How will my information be used?

The UCSF research team will keep your answers confidential and will not share your personal information with anyone outside the research team. Your decision to complete this survey or not, will not impact your medical care at NEMS in any way. Researchers at UCSF will use your information to inform this study. Once the study is completed, we may use de-identified information or share it with other researchers for future research studies. We will not share your name or any other personal information. We will not ask you for additional permission to share this de-identified information.

Are there any risks to me or my privacy?

We will do our best to protect the information we collect from you. We will not collect any information that could identify you, such as your name or address. Authorized representatives from the University of California may review your research data for the purpose of monitoring or managing the conduct of this study.

Are there benefits?

There is no direct benefit to you. Although this does not help you directly, this survey will help us understand people’s testing and treatment preferences and might benefit other people in the future.

Can I say “No”?

Yes, you do not have to participate in this survey. Your decision to complete this survey or not will not impact your medical care at NEMS in any way.

Are there any payments?

When you complete this survey, you will receive a Target gift card of $20 for your participation from UCSF. Please note that it may take the UCSF research team up to 14 business days to send your gift card.

Who can answer my questions about the study?

Please contact Dr. Amy Tang or Dr. Tracy Lin  if you have questions or concerns about your rights as a research participant, you can call the UCSF Institutional Review Board at 415-476-1814.

**Informed consent**

We need to be sure you understood this consent form:

Is your participation in this survey optional?

- Yes
- No

Will your participation in this survey have an impact on the care you receive at NEMS?

- Yes
- No

If you want to participate in this study, check Yes and click the Next button to start the survey.

Do you consent to participate in this study?

- Yes
- No

**Non-consent termination**

You did not consent to participate in this study. This survey will now end.

**Survey introduction Screen**

**Introduction**

Please watch the following video, which contains instructions on how to complete the survey.

**Instruction video script**

**Introduction**

Thank you for agreeing to participate in our survey.

This survey contains 10 different hypothetical, imaginary scenarios that are not real. In each scenario, imagine that your test shows that you have a latent tuberculosis infection. People who have a latent tuberculosis infection are healthy and do not have any symptoms, but they have the tuberculosis germ. This means that they may one day develop active tuberculosis, which is a serious, life-threatening disease. Preventative treatment is available for latent tuberculosis infections. These treatments work by getting rid of the tuberculosis germ. However, if you are travelling to areas where there is high TB risk after preventative treatment, it is possible that you may be reinfected with the tuberculosis germ and contract another latent tuberculosis infection. Preventive treatment is recommended by doctors, but you can decide if you want to take the treatment or not.

Each scenario will describe a preventative treatment for your latent tuberculosis infection that will take 3 to 4 months, with daily or weekly pills. However, the specific characteristics of the preventative treatment will differ between scenarios.

Each scenario of preventative treatment includes the following characteristics:

1. **Risk of developing active tuberculosis without preventative treatment (2-year risk):** This is your chance of developing active tuberculosis in the next two years without preventative treatment. You can still develop active tuberculosis after two years, but the risk is highest in the first two years. If you take medication, your risk will be much lower.
2. **Potential side effects with preventative treatment:** This is a side effect that you may experience. Side effects may include skin rash, fatigue, gastro-intestinal discomfort, liver inflammation, or liver injury which can be reversed by stopping the medication.
3. **Changes to existing medication needed as part of preventative treatment:** The preventative treatment may influence other medicines you may be taking. In some scenarios, you may not have to change anything about your medications. In other scenarios, you may either need to change or stop your current medication.
4. **Out-of-pocket payment cost needed for preventative treatment**: The out-of-pocket cost for healthcare varies. This is what you would have to pay to receive the treatment. You may have to pay $0, $50, or $100.
5. **Clinic visits needed as part of preventative treatment**: Some people need to check their liver and kidney function regularly over the course of the preventative treatment. In some scenarios, you may not need to visit the clinic at all. In other scenarios, you may either need to visit the clinic once or 4 times.
6. **Blood draws needed as part of preventative treatment:** Some people need to have their blood drawn as part of the preventative treatment. In some scenarios, you may not need to have your blood drawn at all. In other scenarios, you may need to have your blood drawn once or have your blood drawn monthly.

After being shown the characteristics of the preventative treatment for your latent tuberculosis infection that is available, you will be asked whether you would accept or decline preventative treatment. There is no right or wrong answer. We are interested in your opinion. Please choose the one you think is more likely, even if you are not sure.

**Comprehension Questions (correct answers italicized and bolded)**

**Comprehension questions**

Before you begin the survey, we would like to confirm that you understood the instructions provided in the video. Please answer the following questions:

1. **How many hypothetical, imaginary scenarios will there be in this survey?**

- 5
- ***10***
- 15

2. **In all of the hypothetical, imaginary scenarios, how long is the preventative treatment being described?**

- 3-4 hours
- 3-4 days
- 3-4 weeks
- ***3-4 months***

3. People **who have a latent tuberculosis infection:**

- Do not have any symptoms
- Have the tuberculosis germ
- Are at risk of developing active tuberculosis
- ***All of the above***

4. **Can you get the** tuberculosis **germ after taking preventative treatment?**

- ***Yes***
- No

**Survey**

Scenario (#)

Imagine your test shows that you have the tuberculosis germ, which means you have a latent tuberculosis infection. Your doctor describes a preventative treatment, which would be 3-4 months long with daily or weekly pills. The characteristics of the preventative treatment are as follows:

| **Characteristic of preventative treatment** | **Possible levels for scenarios** |
| --- | --- |
| **Risk of active TB without preventative treatment** | Low risk  Medium risk  High risk |
| **Potential side effects** | Skin rash  Fatigue  Gastro-intestinal inflammation  Liver injury or inflammation |
| **Changes to existing medication needed** | No change needed  Stop a medicine for 4 months  Switch one of your medicines for 4 months |
| **Out-of-pocket payment cost needed** | $0  $50  $100 |
| **Clinic visits needed** | 0 visits  1 visit  4 visits |
| **Blood draws needed** | No blood draws needed  One blood draw needed  Monthly blood draws needed |

**Given the scenario above, would you accept preventative treatment?**  
(Yes, I would accept preventative treatment/No, I would not accept preventative treatment)

***If yes:* You chose to accept preventative treatment in Scenario (#).**

**If you knew you could be reinfected with the TB germ when travelling to a high TB risk region in the future, what would you do?** 
(I would change and decline preventative treatment/I would still accept preventative treatment)

**Conclusion**

Thank you for participating in this study! If you would like to receive a $20 Target gift card, please click the following link to provide your information:

**Supplemental Figure 1:** Study flow chart. Invitations to the online survey were sent by email and postal mail.

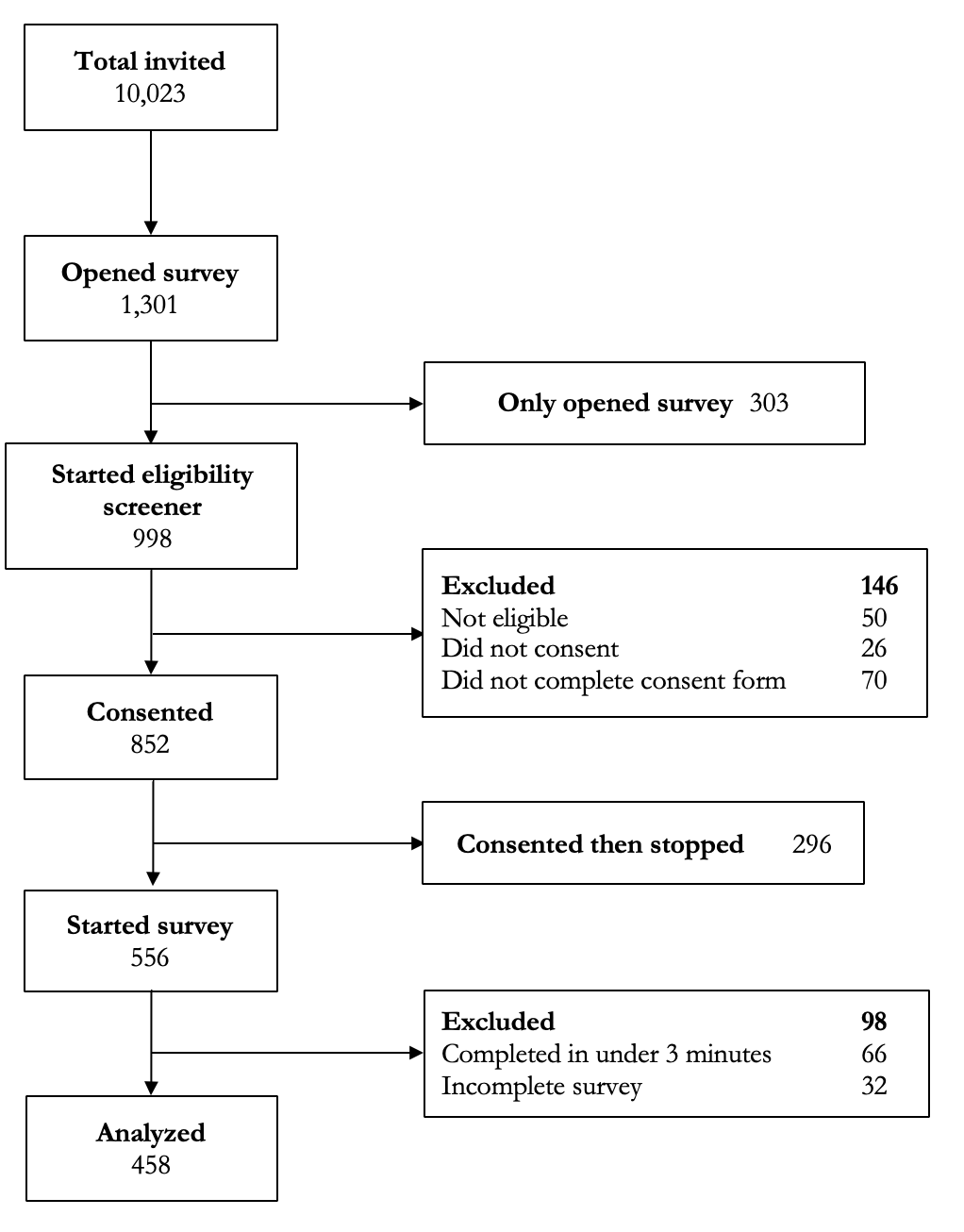

**Supplemental Figure 2:** Sensitivity analysis including reinfection risk (N=458).

This analysis shows the association of all factors with preference for LTBI treatment over no treatment. Adjusted odds ratios (aOR) were calculated using mixed effects logistic regression with fixed effects for all factors and random effects for individual participants. The difference in the predicted proportion of individuals who would prefer LTBI treatment over no treatment was calculated for all combination of scenarios, and we compared the effects of different factor levels (e.g., high vs low risk of tuberculosis) across scenarios. An aOR above 1 means LTBI treatment is more likely to be accepted with this feature (attribute level) compared to the reference.

aOR: adjusted odds ratio, CI: confidence interval, TB: tuberculosis, LTBI: latent tuberculosis infection*,* Δp: difference in predicted proportion accepting LTBI treatment

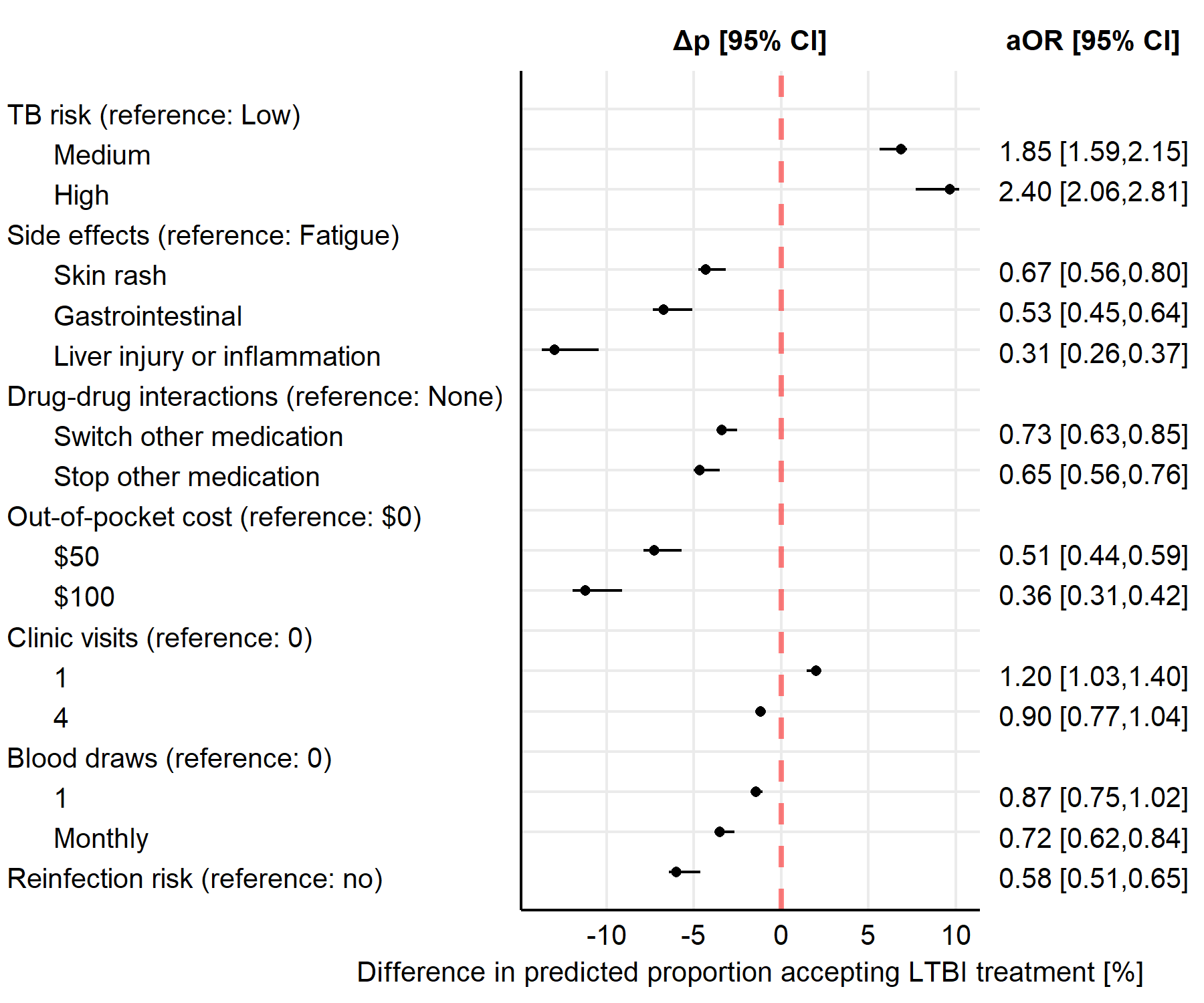

**Supplemental Figure 3:** Sensitivity analysis including partial responses (N=490).

This analysis shows the association of all factors with preference for LTBI treatment over no treatment. Adjusted odds ratios (aOR) were calculated using mixed effects logistic regression with fixed effects for all factors and random effects for individual participants. The difference in the predicted proportion of individuals who would prefer LTBI treatment over no treatment was calculated for all combination of scenarios, and we compared the effects of different factor levels (e.g., high vs low risk of tuberculosis) across scenarios. An aOR above 1 means LTBI treatment is more likely to be accepted with this feature (attribute level) compared to the reference.

aOR: adjusted odds ratio, CI: confidence interval, TB: tuberculosis, LTBI: latent tuberculosis infection*,* Δp: difference in predicted proportion accepting LTBI treatment

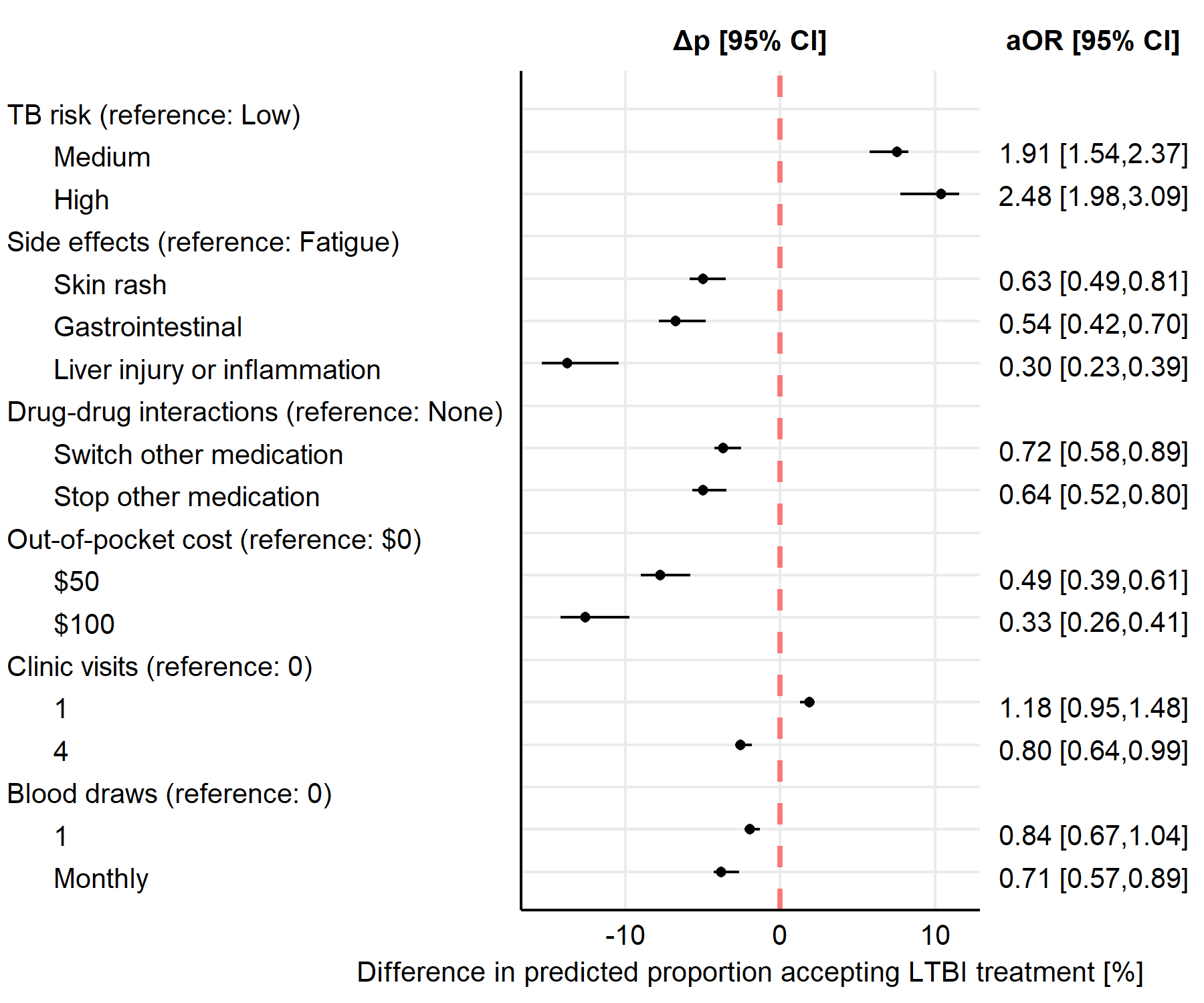

**Supplemental Table 1:** Comparison of age and survey language of individuals invited and responses that were ultimately included in the main analysis. Responses analyzed came from participants who opened the survey, completed the eligibility screener and were eligible, consented, responded to all questions, and completed the survey in 3 or more minutes.

We did not include sex as a variable in the survey and therefore only know the sex distribution for the invited population.

|  | **Individuals Invited** | **Responses Analyzed** |
| --- | --- | --- |
| Total | 10,023 (100%) | 458 (100%) |
| Age |  |  |
| 18-39 | 2,157 (21.5%) | 180 (39.3%) |
| 40-59 | 3,006 (30.0%) | 132 (28.8%) |
| 60+ | 4,860 (48.5%) | 146 (31.9%) |
| Survey language |  |  |
| English | 1,790 (18%) | 58 (12.7%) |
| Traditional Chinese | 2,038 (20%) | 101 (22.1%) |
| Simplified Chinese | 6,195 (62%) | 299 (65.2%) |
| Sex |  |  |
| Female | 6,150 (61.4%) |  |
| Male | 3,873 (38.6%) |  |

**Supplemental Table 2:** Scenarios minimizing and maximizing the predicted acceptance rate, i.e., the predicted proportion of individuals who prefer LTBI treatment over no LTBI treatment.

TB: tuberculosis, LTBI: latent tuberculosis infection

|  | Minimum | Maximum |
| --- | --- | --- |
| TB risk | Low | High |
| Side effects | Liver injury or inflammation | Fatigue |
| Drug-drug interactions | Stop other medication | None |
| Cost | $100 | $0 |
| Clinic visits | 4 | 1 |
| Blood draws | Monthly | 0 |
| Reinfection risk | Yes | No |
| Predicted acceptance rate | **43.0%** | **89.8%** |

**Supplemental Table 3:** Subgroup analysis by age: association of factors with preference for LTBI treatment over no LTBI treatment. The analysis used mixed effects logistic regression (all fixed effects except for random intercept by participant). An adjusted odds ratio (aOR) above 1 means LTBI treatment is more likely to be accepted with this feature (attribute level) compared to the reference. The results are reported for four models: one for each age group, and one for all ages.
aOR: adjusted odds ratio, CI: confidence interval, TB: tuberculosis, LTBI: latent tuberculosis infection

|  | All ages | | Age 18-39 | | Age 40-59 | | Age 60+ | |
| --- | --- | --- | --- | --- | --- | --- | --- | --- |
|  | aOR | [95% CI] | aOR | [95% CI] | aOR | [95% CI] | aOR | [95% CI] |
| TB risk (reference: low) |  |  |  |  |  |  |  |  |
| medium | 1.95 | [1.57, 2.43] | 2.62 | [1.86, 3.69] | 2.11 | [1.37, 3.25] | 1.22 | [0.82, 1.81] |
| high | 2.55 | [2.03, 3.19] | 3.54 | [2.48, 5.05] | 2.18 | [1.39, 3.42] | 2.02 | [1.35, 3.03] |
| Side effects (reference: fatigue) |  |  |  |  |  |  |  |  |
| Skin rash | 0.62 | [0.48, 0.80] | 0.52 | [0.34, 0.78] | 0.53 | [0.32, 0.90] | 0.83 | [0.52, 1.31] |
| Gastrointestinal inflammation | 0.53 | [0.41, 0.69] | 0.40 | [0.26, 0.60] | 0.74 | [0.44, 1.26] | 0.54 | [0.34, 0.85] |
| Liver injury or inflammation | 0.29 | [0.22, 0.38] | 0.27 | [0.18, 0.41] | 0.31 | [0.19, 0.53] | 0.26 | [0.16, 0.42] |
| Drug-drug interactions (reference: none) |  |  |  |  |  |  |  |  |
| Switch other medication | 0.72 | [0.57, 0.90] | 0.73 | [0.52, 1.04] | 0.77 | [0.50, 1.19] | 0.67 | [0.45, 1.00] |
| Stop other medication | 0.64 | [0.51, 0.80] | 0.76 | [0.53, 1.08] | 0.88 | [0.57, 1.37] | 0.43 | [0.29, 0.65] |
| Out-of-pocket cost (reference: $0) |  |  |  |  |  |  |  |  |
| $50 | 0.49 | [0.39, 0.62] | 0.54 | [0.38, 0.78] | 0.60 | [0.39, 0.95] | 0.37 | [0.25, 0.56] |
| $100 | 0.32 | [0.26, 0.41] | 0.35 | [0.25, 0.51] | 0.40 | [0.26, 0.63] | 0.24 | [0.16, 0.36] |
| Clinic visits (reference: 0) |  |  |  |  |  |  |  |  |
| 1 | 1.20 | [0.96, 1.50] | 1.10 | [0.78, 1.55] | 1.13 | [0.72, 1.76] | 1.35 | [0.91, 2.02] |
| 4 | 0.82 | [0.66, 1.03] | 0.81 | [0.57, 1.14] | 0.86 | [0.56, 1.33] | 0.81 | [0.55, 1.21] |
| Blood draws (reference: 0) |  |  |  |  |  |  |  |  |
| 1 | 0.87 | [0.69, 1.08] | 1.03 | [0.73, 1.46] | 0.94 | [0.61, 1.47] | 0.70 | [0.47, 1.04] |
| monthly | 0.75 | [0.60, 0.94] | 0.92 | [0.65, 1.29] | 0.80 | [0.51, 1.24] | 0.57 | [0.38, 0.84] |
